# Estimating the Smallest Worthwhile Difference (SWD) of Psychotherapy for Alcohol Use Disorder: Protocol for a Cross-Sectional Survey

**DOI:** 10.64898/2026.02.16.26346220

**Authors:** Ethan Sahker, Isaac Lu, David Eddie, Ryuhei So, Yan Luo, Kenji Omae, Aran Tajika, James P. Angelo, Toby Crisp, Benjamin Coffin, Toshi A. Furukawa

**Affiliations:** Population Health and Policy Research Unit, Center for Medical Education and Internationalization, Graduate School of Medicine, Kyoto University, Yoshida-Konoe-cho, Sakyo-ku, Kyoto, 606-8501, Japan; Massachusetts General Hospital, Harvard Medical School, 151 Merrimac St, Boston, MA 02114, USA; CureApp, Inc., Kodenma-Cho YS building 4th floor, 12-5 Nihonbashi Kodenma-Cho, Chuo-ku, Tokyo, 103-0001, Japan; Okayama Psychiatric Medical Center, 3-16 Shikata Honmachi, Kita-ku, Okayama City, Okayama, 700-0915, Japan; Department of Health Promotion and Human Behavior, Graduate School of Medicine / School of Public Health, Kyoto University, Yoshida-Konoe-cho, Sakyo-ku, Kyoto, 606-8501, Japan; Scientific Research WorkS Peer Support Group (SRWS-PSG), Osaka, Japan; Department of Next-Generation Organ Transplantation, Graduate School of Medicine, The University of Tokyo, 7-3-1 Hongo, Bunkyo-ku, Tokyo 113-8655, Japan; Department of Innovative Research and Education for Clinicians and Trainees (DiRECT), Fukushima Medical University Hospital, 1 Hikarigaoka, Fukushima City, Fukushima 960-1295, Japan; Senior Counsel, Federal Express Corporation, 1000 FedEx Drive, Moon Township < Pennsylvania 15108; Office of Institutional Advancement and Communications, Kyoto University, Yoshida-Konoe-cho, Sakyo-ku, Kyoto, 606-8501, Japan

**Author notes:** **Correspondence to:** Ethan Sahker, Population Health and Policy Research Unit, Center for Medical Education and Internationalization, Graduate School of Medicine, Kyoto University, Yoshida-Konoe-cho, Sakyo-ku, Kyoto, 606-8501, Japan.

## Abstract

**Background:** Psychotherapy is proven efficacious for the treatment of alcohol use disorder (AUD). However, the patient-perceived importance of its effect is not fully appreciated in the evidence base. The smallest worthwhile difference (SWD) represents the smallest beneficial effect of an intervention that patients deem worthwhile in exchange for the harms, expenses, and inconveniences associated with the intervention, and facilitates the interpretation of patient perceived worthiness of an intervention.

**Methods:** The proposed study will estimate the SWD of NIAAA recommended psychotherapies for AUD treatment with English-speaking American respondents aged 18 and older. Primary participants will be recruited using the Prolific research crowdsourcing site. The SWD will be estimated using the Benefit-Harm Trade-off Method, presenting survey respondents with variable, hypothetical magnitudes of psychotherapy outcomes to find the smallest acceptable effect over a natural recovery alternative. The overall average SWD, and subgroup distributions by participant AUD treatment experiences and AUD symptomology will be described. Secondary findings will estimate the smallest recommendable risk difference for AUD psychotherapy from providers and criminal justice professionals.

**Expected Results:** We expect to find an estimate of the SWD for AUD psychotherapy. Further, we expect that the SWD will vary between clinical subgroups based on AUD symptomology and treatment experiences. We expect differences in SWDs between the general population and those of providers and criminal justice professionals. Findings from this project will inform the treatment decision process about psychotherapy during the clinical consultation for people with AUD.

## INTRODUCTION

Alcohol use disorder (AUD) is one of the most prevalent mental disorders, contributing to significant morbidity and mortality (Global Burden of Disease Collaborative Network, 2024). Effective treatments are available for AUD, but the vast majority of those in need remain untreated (Koob, 2024; Sahker et al., 2023; Tucker et al., 2020). Psychotherapy for AUD is a major option for all treatment seekers. It is supported by a substantial evidence base (Witkiewitz et al., 2019) and is generally well-accepted (NIAAA, 2025). However, psychotherapy can be burdensome to the patient, in terms of expense and inconvenience. Furthermore, most people opt to try to improve without treatment (Fan et al., 2019; Kelly et al., 2017) due to motivating factors such as health, negative personal effects, and costs (Pongsavee et al., 2023; Sobell et al., 2000). Treatment response can be defined as remission (remission from AUD criteria) or recovery (remission from AUD criteria and cessation from high-risk drinking) (NIAAA, 2020). The natural recovery rate (no treatment recovery response rate) is estimated to be about 30% within one year (Burtscheidt et al., 2002; Dawson et al., 2006; Fan et al., 2019; Greene et al., 2023). Importantly, about 20% of people receiving treatment do so through criminal justice pathways (Substance Abuse and Mental Health Services Administration, 2025b), where treatment choices are made by third parties. Key stakeholders involved in the provision and adjudication of mandated treatments need to be involved in optimal treatment selections (Volkow et al., 2017; Winick & Wexler, 2002). Patients, families, providers, criminal justice professionals, and all stakeholders consider many AUD treatment options. The most common option is natural recovery, and psychotherapy is the most common clinical option facilitated by licensed healthcare professionals (Tucker et al., 2020). Considering shared decision-making and patient perspectives, optimal treatment selection must account for both the advantages and disadvantages in relation to available treatment alternatives. However, the patient, provider, and criminal justice professional perspectives on worthwhile treatments for AUD, given the benefits and burdens (i.e., harms, expenses, and inconveniences) have not been evaluated.

The minimal important change (MIC) is one metric used for evaluating the significance of treatment changes in health outcomes. The MIC, also referred to as the minimal important difference or minimal clinically important difference, is defined as the smallest change in health outcome following treatment that is perceived as meaningful by the patient (Jaeschke et al., 1989). It is commonly used to interpret patient-reported outcome measures (PROMs) (Carrasco-Labra et al., 2021). However, the MIC reflects within-person (intra-individual) change from pre- to post-treatment (Hiroe et al., 2005; Leucht et al., 2013), rather than between-person differences. The MIC is limited in several ways: it only applies to a specific outcome assessment tool (Devji et al., 2020; Ferreira et al., 2012), is not connected to any particular intervention (McNamara et al., 2015), and it does not account for the balance between treatment benefits and burdens when compared to other options (Ferreira et al., 2012; McNamara et al., 2015). Therefore, a between-group, treatment-specific, patient-perceived determination of psychotherapy worthiness that accounts for the treatment benefits and burdens is warranted.

The smallest worthwhile difference (SWD) offers an approach to evaluating the perceived importance of an intervention that addresses the shortcomings of the MIC. The SWD is defined as “the smallest beneficial effect of an intervention that justifies the costs, risks, and inconveniences of that intervention over a treatment alternative” (Ferreira et al., 2012). The SWD captures the average patient’s benefit-burden tradeoff when considering two treatment options. It is a patient-centered and intervention-specific measure quantifying the absolute difference in outcomes needed for one treatment to be considered preferable over an alternative (Ferreira et al., 2012; McNamara et al., 2015). The smallest recommendable difference (SRD) uses the same process as the SWD but elicits provider and other professional preferences for treatments. The benefit-harm trade-off method (BHTM) is the standard methodology to estimate the SWD and SRD (Barrett et al., 2005; Sahker et al., 2025; Sahker et al., 2024). The BHTM keeps treatment burdens constant while varying a treatment’s benefits in a clinical scenario. Thus, the BHTM identifies the benefits individuals are willing to accept, given the expected burdens of one intervention over another (Barrett et al., 2005). The BHTM has never been applied to estimate the SWD of AUD psychotherapy.

The present study will estimate the SWD of psychotherapy for AUD based on established methods (Sahker et al., 2024). We will estimate the SWD of recommended AUD psychotherapies (NIAAA, 2025) compared to the natural recovery alternative using the BHTM. We will primarily estimate the SWD for those people who demonstrate moderate-to-severe AUD and have never received AUD treatment (pharmacological or psychosocial). For our secondary analyses, we will examine the SWD of those who are receiving treatment, those with a previous history of AUD, and for those with friends and family with AUD. We will also analyze demographic and clinical predictors of the SWD for exploratory purposes. Additionally, we will estimate the SRD using samples representing psychotherapy providers and criminal justice professionals. The results of this study will assist all stakeholders understand differential patient expectations of psychotherapy and help create an evidence-based benchmark for future clinical trials, recommendations, as well as treatment referrals and mandates.

## METHODS

### Study design

The present study was approved by the Kyoto University Graduate School of Medicine Ethics Committee (R5355), For the primary analysis, we will conduct a cross-sectional survey using Prolific, a research participant crowdsourcing service (RPCS). Recruitment will occur through the Prolific platform, where participants meeting the inclusion criteria for AUD will be directed to an online survey. Prolific represents an approximation to the general population, with participant data demonstrating high test-retest reliability and strong convergent and concurrent validity in psychological research (Chandler & Shapiro, 2016). Eligibility will be restricted to participants residing in the USA. Participants will be drawn from Prolific’s established pool and compensated for their time. Compensation will be $1.50 USD, consistent with remuneration for studies of similar length conducted through RPCSs (Sahker et al., 2024) and based on estimated time-to-completion. Study information will be presented, and informed consent will be obtained electronically (Supplement 1).

We will conduct additional cross-sectional surveys for two secondary analyses with a) psychotherapy providers and b) criminal justice professionals. For psychotherapy providers we will solicit volunteer participation through the American Psychological Association listservs. For criminal justice professionals, we will access the American Bar Association, federal court system, and federal probation listservs, plus emails listed on government websites to recruit judges and probation officers responsible for sentencing and conditions for mandated addiction treatments (Supplement 2).

### Primary outcome and its measurement

The primary outcomes are the smallest worthwhile difference (SWD) and smallest recommendable difference (SRD) of psychotherapy for AUD compared to natural recovery. These represent the smallest difference between psychotherapy and natural recovery deemed worthwhile, given the treatment burdens (harms, expenses, and inconveniences). The measure of treatment effect will be treatment response, defined as AUD recovery (abstinence or asymptomatic low-risk drinking) (NIAAA, 2020; Tucker et al., 2020). We describe this response to participants as “recovery.” Psychotherapy for AUD includes any modality recommended by the National Institute on Alcohol Abuse and Alcoholism (NIAAA): cognitive behavioral therapy (CBT), brief intervention (BI), couples/family therapy, mindfulness-based relapse prevention (MBRP), and dialectical behavioral therapy (DBT) for AUD (Jacobs & Baldessarini, 2002; NIAAA, 2025). The 30% natural recovery rate after one year was estimated from two epidemiological studies both estimating the control event rate (CER) to be 32% (Dawson et al., 2006; Fan et al., 2019), a Cochrane review estimating the CER to be 25% (Greene et al., 2023), and an additional RCT estimating the CER to be 36% (Burtscheidt et al., 2002). Based on the tendency to overestimate the CER in epidemiological studies, and underestimate it in RCTs (Cuijpers et al., 2021; Furukawa et al., 2014), we assume 30% is a reasonable combined estimate.

We will estimate the different participant preferences through the Benefit-Harm Tradeoff Method (BHTM) (Figure 1). The BHTM follows four steps by presenting a clinical scenario and then tradeoff algorithm: 1) we will present a summary of AUD symptoms, 2) we will explain the benefits of the natural recovery alternative to be a 30% response rate within one year (Burtscheidt et al., 2002; Dawson et al., 2006; Fan et al., 2019; Greene et al., 2023), 3) we will explain the potential burdens of NIAAA recommended psychotherapies (Jacobs & Baldessarini, 2002; NIAAA, 2025) in terms of risks (Linden & Schermuly-Haupt, 2014), treatment length (Carey et al., 2008; Cuijpers et al., 2013), outpatient fees (Ark Behavioral Health, 2025; Benson & Song, 2020; Carey et al., 2008; Woods, 2023), and wait times (National Council for Mental Wellbeing, 2022), and 4) we will query if psychotherapy for AUD would be considered worthwhile at varying treatment response rates compared to the natural recovery alternative of 30% response within one year.

**Figure 1.**
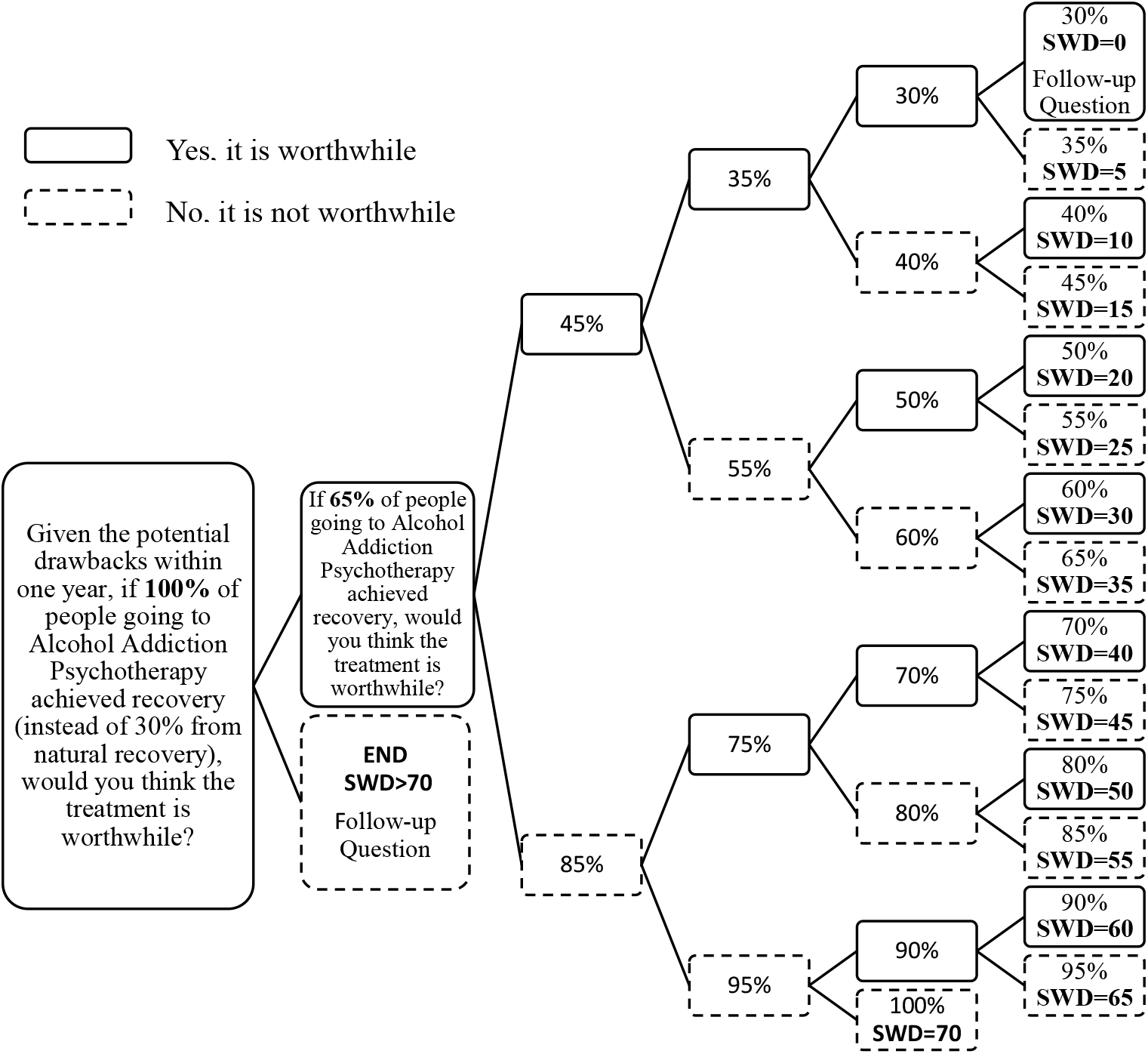
Smallest worthwhile difference (SWD) question sequence algorithm. Participants are queried about their willingness to accept AUD psychotherapy, given the treatment benefits and burdens, until they reach the smallest worthwhile response rate. The first question is repeated but the ratio is replaced by the ratio in the box. The smallest worthwhile response, minus the 30% natural recovery response, equals the individual participant SWD.

In step 4, we will ask participants, “Given the potential drawbacks after one year, if *x*% of people going to psychotherapy recovered (instead of 30% from natural recovery), would you think the treatment is worthwhile?” (Supplement 3). Participants who accept psychotherapy at the variable hypothetical response, given the burdens, will move to a lower response rate. If they reject the response, they will move to a higher response rate. This iterative process will continue until the smallest worthwhile response rate is identified. Finally, the difference between the 30% natural recovery response and the smallest psychotherapy response the participant would consider worthwhile represents the individual participant SWD or SRD.

### Demographic and clinical variables

We will evaluate the current AUD symptom severity using the Alcohol Use Disorders Identification Test (AUDIT) (Saunders et al., 1993). Clinical variables will also include participant lifetime SUD diagnosis, lifetime treatment for SUD (psychosocial or pharmacological), current treatment for SUD, and lifetime non-SUD psychotherapy. In addition, we will collect demographic information including gender, age, race/ethnicity, education, employment, state of residence, and insurance status. Insurance will be categorized as Affordable Care Act, Medicare/Medicaid, private health insurance, Veterans Administration, other, or uninsured. Participant characteristics collected for secondary studies with psychotherapy providers and criminal justice professionals will also include: Credential (in training, licensed counselor, licensed psychologist, certified addictions counselor), Career Level (student, early, mid, late), Type of therapy provided (Addictions, Non-Addictions), criminal justice position (US Attorney, District Court Judge, Circuit Court Judge, Federal Probation Officer, US Marshall, Other), AUD treatment requests (Yes/No), and Preferred treatment (AA/mutual support groups, therapy provided by addictions counselors, therapy provided by clinical psychologists, pharmacotherapies).

After presentation of a clinical scenario and before presentation of the tradeoff algorithm, participants will be asked two attention check questions to gauge performance validity: 1) “What percent of people experience recovery without alcohol addiction/alcohol use disorder treatment? (multiple choice response)”, and 2) “What kinds of burdens are not associated with psychotherapy for alcohol addiction/alcohol use disorder? (multiple choice response)”. After completion of the tradeoff algorithm, participants who reject psychotherapy for AUD at a 100% response rate (burden-sensitive decliners) will be asked, “Why would you decline psychotherapy for alcohol use disorder if the recovery rate was 100%?” ([free entry]). Similarly, participants willing to accept AUD psychotherapy with a 30% recovery rate, equal to natural recovery (burden-tolerant accepters), will be asked, “Why would you accept psychotherapy for alcohol use disorder if the recovery rate was equal to natural recovery?” ([free entry]).

### Clinical Samples

We will include participants from the general population aged 18 or older, living in the USA. The primary sample consists of people exhibiting risky drinking with no history of AUD treatment and are who willing to initiate psychotherapy. This group mimics potential treatment seekers, and we can expect them to provide better estimates of the SWD for AUD as depicted in the provided clinical scenario because of their current experiences and potential treatment needs. This group would represent the best estimate of a clinical sample taken from a general internet population. To further explore how different experiences with AUD and treatment could be associated with treatment-seeking judgements and SWD estimates, we will include participants with four differing profiles:

1. *Risky Drinking but Never Treated*: current moderate-to-severe AUD symptoms (AUDIT ≥ 8) but no lifetime AUD treatment – the primary interest group for SWD estimation
2. *Risky Drinking and Previously Treated*: current moderate-to-severe AUD symptoms (AUDIT ≥ 8) and lifetime treatment history
3. *Low Risk Drinking but Previously Treated*: current subthreshold AUD symptoms (AUDIT < 8) but lifetime treatment history
4. *Low Risk Drinking and Never Treated*: current subthreshold AUD symptoms (AUDIT < 8) and no lifetime treatment for AUD.

### Specialist Samples

We will conduct two secondary studies using the BHTM with specialists that include *Psychotherapy Providers* and *Criminal Justice Professionals*. We will gather additional variables related to education, training, and employment. We will present the clinical scenario to estimate the smallest recommendable difference (SRD). The BHTM for providers will query, “Given the potential drawbacks after one year, if *x*% of people going to psychotherapy experienced recovery (instead of 30% from natural recovery), would you recommend it?” The BHTM for criminal justice professionals will query, “Given the potential drawbacks after one year, if *x*% people going to psychotherapy experienced recovery (instead of 30% from natural recovery), would you request or order it as a mandatory condition of a sentence?” SRD clinical scenarios are provided in Supplement 3B and 3C.

### Sample Sizes

The sample size is set to achieve the expected precision in the estimate of the SWD and SRRD. We assume that the standard deviation (SD) of the SWD would resemble that estimated in our previous study using the same proposed sample with an IQR=10, 35 (approximate SD=20) for SWDs of 20% (Sahker et al., 2024). To obtain a 95% confidence interval (CI) within 10 percentage points, we need approximately 62 participants in each of the four groups described above. Estimates of mental health incidence in RPCSs vary but are demonstrably higher than in the general population (Chandler & Shapiro, 2016) so we may need to recruit fewer people than identified in the general population. Conservative estimates suggest that approximately 9% of the population will comprise the primary interest group *Risky Drinking but Never Treated* (Substance Abuse and Mental Health Services Administration, 2025a), 4% *Risky Drinking and Previously Treated* (Slade et al., 2021), 4.5% *Low Risk Drinking but Previously Treated* (Fan et al., 2019; Slade et al., 2021), and 83% *Low Risk Drinking and Never Treated*. Based on these subgroup population estimates, approximately 1550 participants would be necessary to reach n=62 in all groups and 690 to reach n=62 in the primary interest group (Figure 2). To be sure we reach the needed sample in the group with the smallest occurring incidence, we will aim to recruit between 690 and 1550 participants for the primary study. For the secondary study, we will recruit 100 provider participants, and 100 criminal justice professional participants. In all, 820 to 1800 participants will be collected, depending on response rates in the primary study groups.

**Figure 2.**
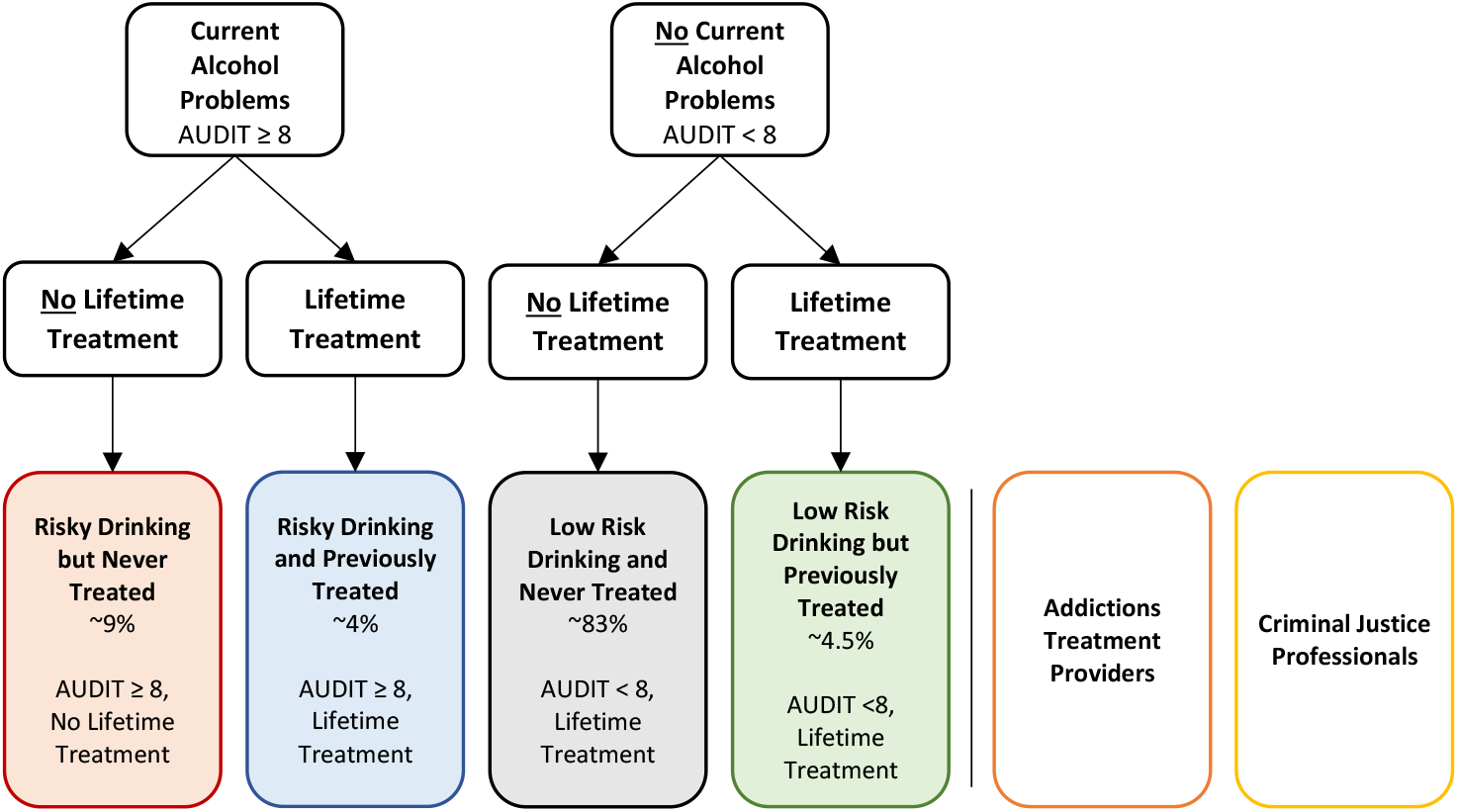
Estimated clinical group representations based on symptomology and treatment experiences. The primary interest being moderate-to-severe depressive symptoms but not in treatment. Red boxes represent subgroup of interest with necessary minimum of n = 62 to calculate the clinical group smallest worthwhile differences, AUDIT = Alcohol Use Disorders Identification Test, % = estimated from general population. Boxes for Addictions Treatment Providers and Criminal Justice Professionals represent sub samples for estimation of the smallest recommendable risk difference.

### Data Analysis

Our primary aim is to present the SWD median and interquartile range (IQR), its distribution, and percentile ranking for people with moderate-to-severe AUD symptoms who have never received SUD treatment (*Risky Drinking but Never Treated*). We will then examine the SWD or SRD for all participant groups for comparison using box-violin plots. We will compare group medians with the Kruskal-Wallis test and post-hoc pairwise Dunn test if significant. We will also conduct three exploratory analyses of the entire sample’s SWD/SRD using demographic and clinical independent variables in a multivariable regression. Model 1 predicts patient SWD from all clinical and demographic variables. Model 2 predicts provider SRD and Model 3 predicts criminal justice professional SRD from all clinical and demographic variables. Regression models will only include responses with no missing outcome data. Participants declining psychotherapy with 100% response (burden-sensitive decliners) will be excluded from the primary analyses, as they are not willing to initiate psychotherapy (McNamara et al., 2015; Sahker et al., 2024).

We will conduct two sensitivity analyses with the primary *Risky Drinking but Never Treated* group: 1) including the burden-sensitive decliners and assigning them an SWD value representing a psychotherapy response rate over 100% (SWD=71), and 2) excluding those with lifetime non-SUD psychotherapy. We will also present descriptive data describing reasons for burden-sensitive decliner and burden-tolerant accepter decisions. SWDs represent a risk difference of the EER between to treatment options with a CER of 30%. SWD and SRDs will be converted to their Cohen’s *d* equivalents for interpretation purposes. We will use R Studio 4.2.2 (R Core Team, 2022) for all statistical analyses.

### Patient and public involvement

The BHTM script has been piloted among people with lived experiences for AUD treatment (coauthors TC and BC) and for criminal justice adjudication (JPA) as part of patient and public involvement (PPI). PPI members reviewed descriptions of AUD based on the DSM-5, benefits and burdens of psychotherapy, and study methods. PPI members provided feedback on survey comprehension and accuracy of the patient, provider, and criminal justice professional experiences. The script and clinical scenarios were modified according to PPI feedback. PPI members will also be invited to collaborate on the analysis interpretation and manuscript write-up.

## DISCUSSION

We expect to estimate the SWD and SRD for AUD psychotherapy for the first time. Further, we expect differential SWD values between clinical groups and specialist groups. Findings from this study will help to inform research and clinical practice by highlighting patient preferences present in shared decision-making practices for AUD psychotherapies.

Considering interpretation of our findings, the existing CER and experimental event rate (EER) for AUD need to be identified in the literature. For the CER, natural recovery will represent the control comparison treatment alternative. For the EER, recommended psychotherapies for AUD represent the primary treatment option, which is the target of the SWD. The EER is altered in the BHTM to determine participants’ smallest acceptable treatment effect compared to the CER. However, the currently available EER of psychotherapy for AUD is necessary for interpreting clinical importance.

The natural recovery, or no treatment, response rate for AUD for the present study is estimated to be 30% within one year. This response rate is estimated based on the combination of clinical trials and epidemiological studies. However, the true response rate experienced by patients not receiving treatment is difficult to estimate. For example, the control response rates from clinical trials tend to be underestimated because control participants know they are not receiving the treatment they want leading to worsening symptoms (Furukawa et al., 2014). Among all psychotherapy trials, waitlist controls tend to have lower response rates than usual care controls (17% vs. 20-30%, respectively) (Cuijpers et al., 2021). Control comparisons also demonstrate differential effects in CBT for AUD studies comparing no treatment (often waitlist), attention-placebo, and usual care control conditions (Magill et al., 2023). Differential recovery rates in control comparisons are due to many methodological differences, and control group type is clearly part of the reason (Magill et al., 2019, 2023; Tucker et al., 2020). Alternatively, evidence from non-controlled epidemiological studies can overestimate the true natural recovery rate, due to self-reporting biases, and a lack of clinical diagnostics (Sobell et al., 2000). Yet, the best approximation for the true natural recovery rate comes from epidemiological results. These studies show that about 32% respond without treatment within the year (Dawson et al., 2006; Fan et al., 2019). This estimate is supported by a meta-analysis with an expected smaller CER of 25% (Greene et al., 2023), and a clinical trial with an expected greater CER of 36%, using usual care support groups as a control (Burtscheidt et al., 2002). Considering variability and potential overestimation, we have determined that a 30% response rate is a reasonable estimate.

The psychotherapy response rate for AUD is estimated to be 40%. This comes from the summary evidence presented in meta-analyses and epidemiological findings. Determining the response rate with recovery as an outcome is difficult to determine from the literature. One very comprehensive meta-analysis (Magill et al., 2019) included 14 RCTs of psychotherapies for AUD, but only one study measured abstinence rate, which can be interpreted as recovery. They found that 36% of the treatment group recovered within one year (Burtscheidt et al., 2002). Recovery could be assumed to be greater than 36%, given that more participants moderated drinking below diagnostic recovery levels, but did not achieve abstinence. Epidemiological studies report EERs with 40% (Fan et al., 2019) and 46% (Dawson et al., 2006). However, these treatment response rates include all types of treatment and are dominated by self-help group attendance. Trials measuring recovery as an outcome with recommended psychotherapies for AUD are rare. However, a meta-analysis included seven trials implementing recommended psychotherapies for AUD versus usual care or wait list and found that the psychosocial treatments to be superior (risk ratio=1.44 95% CI=1.19, 1.76), which was calculated with a 39% EER (Greene et al., 2023). Additionally, another RCT using CBT-based coping skills training compared to support groups demonstrated a 40% abstinence response rate (Burtscheidt et al., 2002). Taken together the response rate can be assumed to be somewhere between 39-45%. It is safe to estimate a conservative 40% response rate within a year, given the combined evidence and the assumption recovery (abstinence or asymptomatic with low-risk drinking) is greater than the abstinence response rate.

## Data Availability

All data produced in the present study are available upon reasonable request to the authors.

## ACKNOWLEDGMENTS

ES is supported by the Japan Society for the Promotion of Science (grant 24K20239).

## ETHICAL APPROVAL

This research has been approved by the Kyoto University Graduate School of Medicine, Faculty of Medicine, and Hospital Medical Ethics Committee (R5355). All participants will provide e-consent.

