## Supplement for "Estimating the Smallest Worthwhile Difference (SWD) of Psychotherapy for Alcohol Use Disorder: Protocol for a Cross-Sectional Survey"

### Estimating the Smallest Worthwhile Difference (SWD) of Recommended Psychotherapies for Alcohol Use Disorder: An Observational Study

#### Article Supplement

**Supplement 1.** Study questionnaire

**Supplement 2.** Recruitment invitation email for treatment provider and criminal justice professional samples

**Supplement 3.** Clinical scenarios for the smallest worthwhile difference (SWD), smallest recommendable risk difference (SRD), and phrasing for the benefit harm tradeoff method (BHTM).

**Supplement 4.** Response rate to Cohen's *d* estimation method

**Supplement 1.** Study questionnaire

*** SCREENING for Psychotherapy Providers
**Do you provide psychotherapy of any kind?**

- Yes [screen in]
- No [screen out] ***

**Electronic Description Document**

**Study Title:** Estimating the Smallest Worthwhile Difference (SWD) of Psychotherapy for Alcohol Use Disorder

**Research Summary**:
***Ethics review and permission***: Prior to onset, this study was reviewed and approved by the Medical Ethics Committee of the Kyoto University Graduate School of Medicine and Faculty of Medicine, the Kyoto University Hospital, and the head of the research institute.

***Name of research institute and name of principal investigator***: We are researchers at Kyoto University, led by Dr. Ethan Sahker, PhD, in collaboration with researchers at Massachusetts General Hospital / Harvard Medical School (David Eddie, PhD).

***Reason for being selected as a research subject***: You were selected for this survey because you are representative of the general population aged 18 or older, provide psychotherapy, or adjudicate sentences mandating addiction treatment.

***Study objectives and significance***: We are studying patient preferences in the treatment of depression. The purpose of this study is to determine how to evaluate the effectiveness of psychotherapy from the patient's perspective, considering the benefits and disadvantages of psychotherapy. We believe that the survey results will help healthcare professionals and researchers understand patient expectations regarding the effectiveness of psychotherapy and accurately interpret the importance of the results obtained from clinical research.

***Study method***: We plan to recruit approximately 1800 participants. Those who agree to participate will be asked to complete an online survey that takes approximately 5 to 10 minutes. First, participants will be asked about their mental health and past treatment history. Next, we will explain depression and the effectiveness of psychotherapy, as well as the burdens and costs. We will then ask your opinions on the effectiveness of psychotherapy through a series of questions.

***Duration of the study***: From the date of approval by the head of the research institution to September 30, 2028.

**Risks and Benefits**: Risks to participants are expected to be minimal. There is a time commitment to complete the questionnaire. In addition, collecting data over the Internet and normal Internet use are thought to be equally strenuous. There is no financial burden on participants. Participation in this study may not directly benefit participants, but it provides an opportunity to gain knowledge about depression, as well as the costs and benefits of psychotherapy. It will also contribute to important issues such as how results obtained in future clinical studies could be interpreted to incorporate patient values into clinical trials.

**Financial burden and compensation for research subjects:** In principle, there is no financial burden associated with participating in this study, but participants will be responsible for communication costs associated with connecting to the Internet when answering the questionnaire. There is no compensation for treatment providers and criminal justice adjudicators and is completely volunteer. Participants recruited through Prolific will be compensated for the time and effort spent answering the questionnaire in accordance with the terms of Prolific.

**Handling of personal information:** Participant IDs will be managed in accordance with Prolific's Participant Agreement and Privacy Policy. Only the principal investigator will have access to the website ID, but it will be used to allocate rewards and will not be shared with others. Survey data will be stored on the secure servers of the crowdsourcing websites, but they will not have access to survey data. Data will be backed up by the principal investigator. After data collection is complete, all IDs will be deleted, and data will be deidentified to make it unidentifiable. Once the survey is complete, all data will be deleted from websites' servers. Deidentified data will be stored on a password-protected local server in the principal investigator's lockable room for 10 years after publication of the primary results. Thereafter, the data stored so that personal information cannot be known and identifying information will be destroyed. No personally identifiable information will be included in any papers or conference presentations related to the results of this study. Secondary use in other research and provision to other research institutions will not include any personally identifiable information.

**When providing information to a person in a foreign country:**

1. Universities: Harvard University
2. Information on the system for protecting personal information in the foreign country obtained in an appropriate and reasonable manner:
   Detail: https://www.ppc.go.jp/personalinfo/legal/kaiseihogohou/#gaikoku
   - Electronic Communications Privacy Act of 1986 (ECPA)
     - A. URL: https://bja.ojp.gov/program/it/privacy-civil-liberties/authorities/statutes/1285
     - B. Status of Implementation: Effective October 21, 1986
     - C. Covered Entities: Public sector entities (including local governments) and private entities that electronically store personal data1
     - D. Covered Information: "Electronic Communications" (transmission of symbols, signals, text, images, sounds, data, or information of any nature, transmitted in whole or in part by wire or electronic system)
   - Gramm Leach Bliley Act (GLBA)
     - URL: https://www.ftc.gov/tips-advice/business-center/privacy-and-security/grammleach-bliley-act
     - Status of Implementation: Effective November 12, 1999
     - Target institutions: Private financial institutions "significantly engaged" in the financial services industry
     - Target information: "Non-Public Personal Information" (all information collected from customers through the provision of financial services)
3. Information on the measures taken by the person to protect personal information: ID management for Prolific. Only the Principal Investigator has access to the participant ID, but it will not be shared with anyone other than the Principal Investigator and will be used only for the purpose of distributing rewards. Survey data will be stored on the secure servers of Prolific, but this company will not have access to the survey responses. Once data collection is complete, participant IDs will be removed from the dataset and the data will be anonymized so that individuals cannot be identified. The anonymized data will be stored in the principal investigator's locked room on a password-protected local server for 10 years after the publication of the main results. The data will then be erased so that personal information cannot be identified, and the media will be disposed of. The anonymized data will be shared with collaborating universities, but no information will be linked to participants. Reports and presentations on the findings of this study will not contain any personally identifiable information.

**Potential for secondary use of information or provision to other research institutions:** Information collected in this study may be used for future research that is not specified at the time of obtaining consent. When used for secondary research or provided to other research institutions, no personally identifiable information will be included. If you would like to confirm future research, you can contact the principal investigator (see contact information below).

**Voluntary Participation**: Participation in this study is voluntary. Participants may withdraw consent at any time during the survey without giving a reason. Those withdrawing will not be disadvantaged in any way if they do not agree to participate or if consent is withdrawn. If consent is withdrawn, we will not use the data collected. However, once the survey is completed, your name is not connected to the data and delete the data in not possible. Materials related to this research may be obtained and viewed to the extent that it does not interfere with the personal information and intellectual property of other research subjects and others. The decision to participate or not to participate will not affect current or future relationships with Kyoto University or Prolific.

**Research Funding and Conflicts of Interest**: This research is funded by the Kyoto University Graduate School of Medicine. Conflicts of interest are appropriately reviewed by the Kyoto University Clinical Research Conflict of Interest Review Committee in accordance with the Kyoto University Conflict of Interest Policy and the Kyoto University Conflict of Interest Management Regulations.

**Contact Information**: For further information, please contact Dr. Ethan Sahker, Junior Associate Professor, Population Health and Policy Research Unit, Center for Medical Education and Internationalization, Graduate School of Medicine, Kyoto University:.
Kyoto University Contact: Research Promotion Division, General Affairs and Planning Department, Graduate School of Medicine, Kyoto University


**By selecting "I agree" to the first question on the survey form you agree to participate in this study. Please also read your crowdsourcing website's Participant Agreement and Privacy Agreement as they pertain to the management of individual website IDs. Have you read the instructions and agree to participate in this study?**

- I agree
- I do not agree

**What is your current age? _________**

**What is your sex assigned at birth?**

- Male
- Female

**What race/ethnicity do you identify as most?**

- White or Caucasian
- Black or African American
- Latino or Hispanic
- Asian
- Indian (Asian Indian Subcontinent)
- Middle Eastern, Arab, or Persian
- Native American or Alaska Native
- Native Hawaiian or Pacific Islander
- Multiracial
- Other

**What is the highest level of education you have completed?**

- Less than high school
- High school graduate / GED
- Some college
- 2-year degree
- 4-year degree
- Master's Degree
- Doctorate (PhD, MD, JD, etc.)

**What best describes your current employment status?**

- Working full-time
- Working part-time

- Unemployed and looking for work
- Homemaker or stay-at-home parent
- Student
- Retired
- Disabled

**In which State do you reside? -** List 50 states, DC, and Puerto Rico

**Which best describes your health insurance status?**

- Medicare / Medicaid
- VA Benefits / TRICARE
- Affordable Care Act / Obamacare
- Private Health Insurance
- Uninsured

**Are you currently receiving treatment for a drug or alcohol addiction (Therapy, Group, Alcoholics Anonymous, medications, etc.)?**

- Yes
- No

**Have you ever received treatment for a drug or alcohol addiction (Therapy, Group, Alcoholics Anonymous, medications, etc.)?**

- Yes
- No

**Have you ever received any psychotherapy or counseling for other mental health concerns (depression, anxiety, etc.)?**

- Yes
- No

**Have you ever been diagnosed with alcohol use disorder by a medical or mental health professional?**

- Yes
- No

**Have you ever felt that you may have an alcohol addiction or alcohol problem?**

- Yes
- No

**Have you ever been diagnosed with any other substance use disorder by a medical or mental health professional?**

- Yes
- No

**Have you ever felt that you may have a drug addiction or drug problem (other than alcohol)?**

- Yes
- No

**Please answer the next questions about alcohol based on the Alcohol Unit Reference image representing 1 standard drink for different types of alcohol.**

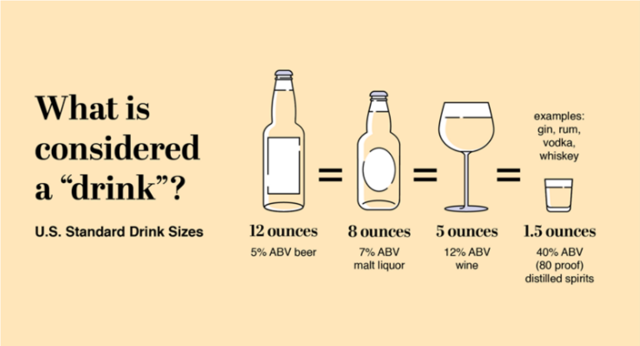


| | | | | | |
|---|---|---|---|---|---|
| How often do you have a drink containing alcohol? | Never | Monthly or less | 2 to 4 times per month | 2 to 3 times per week | 4 times or more per week |
| How many drinks containing alcohol do you have on a typical day when you are drinking? | 0 to 2 | 3 to 4 | 5 to 6 | 7 to 9 | 10 or more |
| How often do you have six or more drinks on one occasion? | Never | Less than monthly | Monthly | Weekly | Daily or almost daily |
| How often during the last year have you found that you were not able to stop drinking once you had started? | Never | Less than monthly | Monthly | Weekly | Daily or almost daily |
| How often during the last year have you failed to do what was normally expected of you because of drinking? | Never | Less than monthly | Monthly | Weekly | Daily or almost daily |
| How often during the last year have you needed a first drink in the morning to get yourself going after a heavy drinking session? | Never | Less than monthly | Monthly | Weekly | Daily or almost daily |
| How often during the last year have you had a feeling of guilt or remorse after drinking? | Never | Less than monthly | Monthly | Weekly | Daily or almost daily |
| How often during the last year have you been unable to remember what happened the night before because of your drinking? | Never | Less than monthly | Monthly | Weekly | Daily or almost daily |
| Have you or someone else been injured because of your drinking? | No | | Yes, but not in the last year | | Yes, during the last year |
| Has a relative, friend, doctor, or other health care worker been concerned about your drinking or suggested you cut down? | No | | Yes, but not in the last year | | Yes, during the last year |

**What would be your treatment preference for substance use disorders or addiction?**

- Psychotherapy (Counseling: Group or Individual)
- Support Groups (Alcoholics Anonymous, Rational Recovery, 12-Step Groups, Mutual Aid)
- No preference

**[ADDITIONAL QUESTIONS FOR PROVIDERS]**

**What is your relevant psychotherapy credential?**

- In Supervised Training
- Certified Addictions Counselor
- Licensed Professional Counselor / Licensed Mental Health Counselor

- Licensed Marriage and Family Therapist
- Licensed Clinical Social Worker
- Licensed Psychologist

**What is your career level?**

- Student
- Early Career (0-10 years post degree)
- Mid-Career (11-20 years)
- Late-Career (21+ years)

**Do you provide treatment for substance use disorders?**

- Never
- Rarely
- Occasionally
- Frequently
- Primarily

**[ADDITIONAL QUESTIONS FOR CRIMINAL JUSTICE PROFESSIONALS]**

**What is your position in the criminal justice system?**

- US Attorney
- District Court Judge
- Circuit Court Judge
- Federal Probation Officer
- US Marshall
- Other [free text]

**What is your career level?**

- Early Career (0-10 years post degree)
- Mid-Career (11-20 years)
- Late-Career (21+ years)

**Do you make requests or orders for drug and alcohol treatment as part of your duties?**

- Never
- Rarely
- Occasionally
- Frequently
- Primarily

**-- PRESENT BHTM CLINICAL SCENARIO --**

**[AFTER CLINICAL SCENARIO]**

**What percent of people experience recovery from alcohol addiction/alcohol use disorder without treatment?**

- 0%
- 10%
- 20%
- 30%
- 40%

**What kinds of burdens are NOT associated with psychotherapy for alcohol addiction/alcohol use disorder?**

- Side Effects
- Scheduling Difficulties
- Travel Expenses
- Treatment Costs
- Time Burdens

**[AFTER SURVEY COMPLETION]**

**Why would you decline psychotherapy for alcohol addiction/alcohol use disorder if the recovery rate were 100%?**

- Other [free entry]

**Why would you accept psychotherapy for alcohol addiction/alcohol use disorder if the recovery rate were 30%, which is equal to Natural Recovery?"**

- Other [free entry]

**Supplement 2.** Recruitment invitation email for treatment provider and criminal justice professional samples

**Subject:**
Invitation to Participate in Research on [Provider / Criminal Justice] Perspectives on Addiction Treatment Efficacy

**Body:**
Dear Colleague,

My name is Ethan Sahker, PhD, and I am a researcher from Kyoto University. We are conducting a study titled *Estimating the Smallest Worthwhile Difference (SWD) of Psychotherapy for Alcohol Use Disorder*, and I am writing to invite eligible individuals to participate. This study has been reviewed and approved by the Graduate School of Medicine Ethics Committee at Kyoto University, approval number: R5355.

**Purpose of the Study**
The purpose of this research is to determine the smallest treatment effect of addition psychotherapy that [treatment providers / criminal justice professionals] perceive as worthwhile for client referrals, given the treatment risks, expenses, and inconveniences.

**Who Can Participate**
You may be eligible to participate if:

[Providers]

- You are a psychotherapy provider
- Provide or refer clients to specialty addiction treatment

[CJ Professionals]

- You are a Judge, attorney, probation officer, parole officer
- You request or order addiction treatment as a mandatory condition of a sentence

**What Participation Involves**
If you choose to participate, you will be asked to complete an online survey that will take ***7 minutes on average***. Participation is completely voluntary, and you may withdraw at any time without penalty.

**Potential Risks and Benefits**
Risks are minimal and may include emotional reflection. There are no direct benefits to you, but your participation may contribute to research that can improve Your perspective as a [treatment provider / criminal justice professional] is vital to understanding efficacy benchmarks for recommendable therapies and clinical trial development.

**Confidentiality**
All information you provide will be kept confidential. Data will be stored securely and reported without identifying information. Only the research team will have access to the data.

**Compensation**
There is ***no compensation*** and participation is on a volunteer basis.

**Participation**

If you are interested, please click the link below to learn more and begin the survey:
[Insert survey link]

Thank you for considering this invitation! We greatly appreciate your time and contribution to advancing research on substance use treatment. If you have any questions, please contact the Principal Investigator at

Warm regards,
Ethan

**Ethan Sahker, PhD**
Junior Associate Professor
Population Health and Policy Research Unit
Center for Medical Education and Internationalization
Graduate School of Medicine
Kyoto University
T: +81-75-753-9323


**Supplement 3.** Clinical scenarios for the smallest worthwhile difference (SWD), smallest recommendable risk difference (SRD), and phrasing for the benefit harm tradeoff method (BHTM).

**[Page 1] ------------------------------**
**The next five slides include information on Alcohol Use Disorder (alcohol addiction) and its treatment. Please carefully read them and keep this in mind when answering the questions after the information.**

**ALCOHOL USE DISORDER (ALCOHOL ADDICTION)**
Many people drink alcohol from time to time. For some, drinking becomes risky or hard to control and starts to interfere with daily life. When this pattern persists and causes significant impairment or distress, it may reflect Alcohol Use Disorder (AUD), a diagnosable psychiatric condition that lasts at least 12 months and may recur or persist over time. To be diagnosed with Alcohol Use Disorder, symptoms such as the following must be present:

- Alcohol is often taken in larger amounts or over a longer period than was intended.
- There is a persistent desire or unsuccessful efforts to reduce or control alcohol use.
- A great deal of time is spent in activities necessary to obtain alcohol, use alcohol, or recover from its effects.
- Craving, or a strong desire or urge to use alcohol.
- Recurrent alcohol use resulting in a failure to fulfill major role obligations at work, school, or home.
- Continued alcohol use despite having persistent or recurrent social or interpersonal problems caused or exacerbated by the effects of alcohol.
- Important social, occupational, or recreational activities are given up or reduced because of alcohol use.
- Recurrent alcohol use in situations in which it is physically hazardous.
- Alcohol use is continued despite knowledge of having a persistent or recurrent physical or psychological problem that is likely to have been caused or exacerbated by alcohol.
- Tolerance, as defined by either:
  - A need for markedly increased amounts of alcohol to achieve intoxication or desired effect, or
  - A markedly diminished effect with continued use of the same amount of alcohol.
- Withdrawal, as manifested by either:
  - The characteristic withdrawal syndrome for alcohol, or
  - Alcohol (or a closely related substance, such as a benzodiazepine) is taken to relieve or avoid withdrawal symptoms.

Not everyone with Alcohol Use Disorder experiences every symptom. Some people experience only a few symptoms, while others may experience many. Symptoms are usually severe enough to cause noticeable problems with relationships, work, school, at social activities.

**[Page 2] ------------------------------**
**Please carefully read the following and keep this in mind when answering the remaining questions.**

**TREATMENT OF ALCOHOL USE DISORDER (ALCOHOL ADDICTION)**
Recovery from Alcohol Use Disorder means that the symptoms are eliminated, and high-risk drinking has stopped. There are many different approaches to treating Alcohol Use Disorder, but in this study, we will only focus on two options, (1) Natural Recovery and (2) Alcohol Use Disorder Psychotherapies (i.e., psychological intervention, counseling, talk therapy). Alcoholics Anonymous is not a psychotherapy. First, I will describe each treatment option's expected benefits and drawbacks. Then, I will ask if you think Alcohol Use Disorder Psychotherapy is worthwhile at different recovery response rates.

**(1) No Treatment (Natural Recovery)**
Without treatment, about 30% of people can expect to achieve recovery within one year.

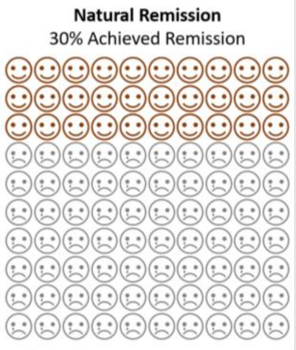

**[Page 3] ------------------------------**
**Please carefully read the following and keep this in mind when answering the remaining questions.**

**<u>TREATMENT OF ALCOHOL USE DISORDER (ALCOHOL ADDICTION)</u>**

**(2) Psychotherapy for Alcohol Use Disorder (i.e., psychological intervention, counseling, talk therapy, group)**

Psychotherapy for Alcohol Use Disorder includes a range of counseling or psychological interventions used to treat clinical symptoms. There are many different types of psychotherapies and formats backed by evidence which are equally effective. For example, individual face-to-face or group cognitive behavioral therapy (CBT) is one recommended psychotherapy.

There are no side effects associated with psychotherapy. However, there are burdens that patients experience:

- Treatment Cost Range = $0-$350 (depending on program and location)
    - Average insured session = $21 and court
    - Average Court mandated = $23 per session (not including court and administrative costs)
- Travel Expenses
- Scheduling Difficulties
- Time spent in therapy (11 weekly 50-minute sessions on average)
- Time spent on treatment homework
- Court mandated treatment varies by treatment level and location, but Level 1 outpatient can last about 20 weeks.
- Wait for treatment is 48 days on average, but this varies on location.

**[Page 4] ------------------------------**
**Please carefully read the following and keep this in mind when answering the remaining questions.**

Remember to only consider two treatment options: (1) Natural Recovery and (2) Alcohol Use Disorder psychotherapies (psychological intervention, counseling, talk therapy). Do not consider any other alternatives like Alcoholics Anonymous in your decision.

When answering, first consider the possible drawbacks we've mentioned (expenses and other inconveniences). Then, weigh the drawbacks against the presented hypothetical benefits when answering the following questions.

**A. <u>TRADEOFF QUESTIONS</u> – General Population [Page 5+]**

Given the potential drawbacks after 1 year, if 100% of people going to Psychotherapy for Alcohol Use Disorder achieved recovery (instead of 30% from Natural Recovery), would you think the treatment is worthwhile?

**B. <u>TRADEOFF QUESTIONS</u> – Providers [Page 5+]**

Given the potential drawbacks after 1 year, if 100% of people going to Psychotherapy for Alcohol Use Disorder achieved recovery (instead of 30% from Natural Recovery), would you recommend it?

**C. <u>TRADEOFF QUESTIONS</u> – Criminal Justice Professionals [Page 5+]**

Given the potential drawbacks after 1 year, if 100% of people going to psychotherapy for Alcohol Use Disorder achieved recovery (instead of 30% from Natural Recovery), would you recommend, request, or order it as a mandatory condition of a sentence?

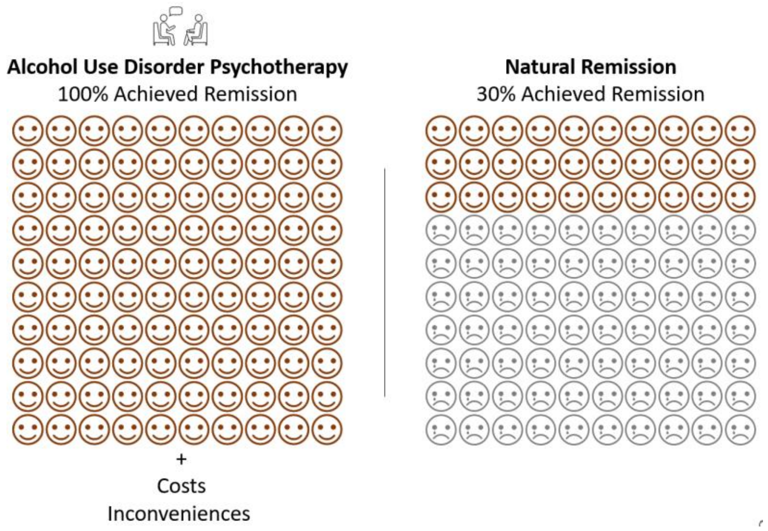


***Respondent responses follow an algorithm presented in Figure 1 in the main text, altering the response proportion/ratio and the representative psychotherapy face figure.***

**Supplement 4.** Experimental event rate to Cohen's *d* estimation method.

Computing Cohen's d for an experimental event rate (EER) of 45% and a control event rate (CER) of 30% using the log odds ratio (log OR) method (Chinn, 2000):

#### Formula for Cohen's *d* from Odds Ratio:

1. Compute the Odds Ratio (OR):

$$OR = \frac{p_1\,/(1 - p_1)}{p_0\,/(1 - p_0)}$$

   where:

   - $p_1$=0.45 (treatment response rate)
   - $p_0$=0.30 (control response rate)

2. Convert OR to Cohen's d:

$$d = \frac{\log{(OR)}}{1.81}$$

   given:

   - the standard deviation of the log odds distribution (logistic model) is:

$$\frac{\pi}{\sqrt{3}} \approx 1.81$$

**R Script:**

```
# Given response rates
p1 <- 0.45  # Experimental Event Rate
p0 <- 0.30 # Control Event Rate

# Compute Odds Ratio (OR)
OR <- (p1 / (1 - p1)) / (p0 / (1 - p0))

# Compute Cohen's d
d <- log(OR) / 1.81

# Print result
print(d)
```

Chinn S. A simple method for converting an odds ratio to effect size for use in meta-analysis. Stat Med 2000; 19: 3127–31.
